# How Many COVID-19 Patients Will Need Ventilators Tomorrow?*

**DOI:** 10.1101/2020.05.18.20105783

**Authors:** J. G. Dai, Mark Gluzman, Alyf Janmohamed, Yaosheng Xu

## Abstract

This paper develops an algorithm to predict the number of Covid-19 patients who will start to use ventilators tomorrow. This algorithm is intended to be utilized by a large hospital or a group of coordinated hospitals at the end of each day (e.g. 8pm) when the current number of non-ventilated Covid-19 patients and the predicated number of Covid-19 admissions for tomorrow are available. The predicted number of new admissions can be replaced by the numbers of Covid-19 admissions in the previous *d* days (including today) for some integer *d* ≥ 1 when such data is available. In our simulation model that is calibrated with New York City’s Covid-19 data, our predictions have consistently provided reliable estimates of the number of the ventilator-starts next day. This algorithm has been implemented through a web interface at `covidvent.github.io`, which is available for public usage.

Utilizing this algorithm, our paper also suggests a ventilator ordering and returning policy. The policy will dictate at the end of each day how many ventilators should be ordered tonight from a central stockpile so that they will arrive by tomorrow morning and how many ventilators should be returned tomorrow morning to the central stockpile. In 100 runs of operating our ventilator order and return policy, no patients were denied of ventilation and there was no excessive inventory of ventilators kept at hospitals.

## 1 Predicting number of new patients on vents

Throughout this paper, the term “patient” refers to Covid-19 patient and the term “hospital” refers to one large hospital that treats Covid-19 patients or a group of hospitals in a region that have some central coordination on ventilators. Let S be the number of new patients who will require ventilator support next day (tomorrow) at this hospital. We will call these patients “vent-starts” or “vent-start patients”. We provide the estimate of the number of ‘vent-starts’ at the end of each day when the hospital’s daily numbers of on-vent and not-on-vent patients become available. We estimate this quantity using the dynamic data (available today from the hospital) and parameters (available historically, not necessarily from the hospital) as described below.

### Dynamic data

The following dynamic data can be observed or estimated daily.

(D.1) *L*: the number of hospitalized patients who are not on vents today.
(D.2) *A*: an estimate of the next day’s number of new hospital admissions. We consider a time-series method for predicting *A* in Section 4.4.

### Parameters

The following parameters are static and can be estimated from historical data.

(P.1) The probability for a newly admitted patient to belong to one of these types
  type 1: never-vent patient; patient will never use a vent.
  type 2: vent-cure patient; patent will use a vent and be cured. type 3: vent-die patient; patient will use a vent and die later.
  These probabilities are denoted by

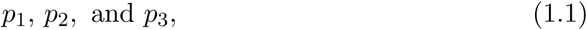

respectively. We assume that each hospitalized patient belongs to one of the these three types even though the type is not observable at admission time.
(P.2) The average length-of-stay (LOS)

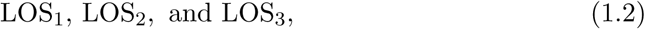

for each type of patient. Specifically,
  a. For type 1 (never-vent), the length-of-stay is equal to the number of days in hospitalization, from admission to discharge.
  b. For type 2 (vent-cure) and type 3 (vent-die) patients, the length-of-stay is equal to number of days in hospitalization *before* ventilation, from admission to ventilation. Therefore, for a type 2 or type 3 patient, the length-of-stay can be better called time-to-ventilation.

In this paper we argue that the expected number of vent-starts next day can be estimated as

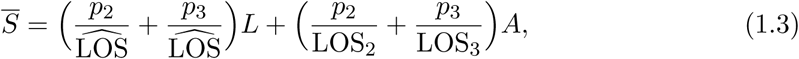

where

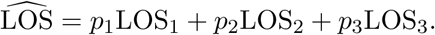

Of course, the number of vent-starts next day *S* is random. We will argue that *S* can be modeled as a random variable that follows Poisson distribution with mean 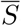:

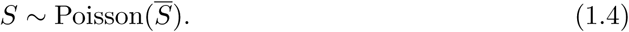

Using this model, one can easily compute an upper confidence interval bound *U* that satisfies

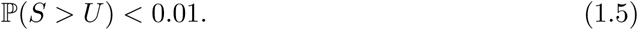

In Section 3, we will use the upper confidence bound *U* to design a policy for ordering and returning ventilators.

## 2 Methodologies to justify (1.4)

Suppose that we are at the end of day *t*. We identify two groups of patients who potentially can become vent-start patients on day *t* + 1: hospitalized patients who are not on the vent support (not-on-vent patients) on day *t* and new patients who will be admitted on day *t* + 1.

We denote *L_t_* as the number of non-ventilated patients at the end of day *t* who might need the vent support in the future. (We assume *L_t_* does not include non-ventilated patients who have previously been on vent support.) On day *t*, there are *L_t_* not-on-vent patients in hospitals; a fraction of them, 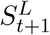, will turn into vent-patients on day *t* + 1.

We assume that the number of vent-start patients 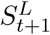 should be proportional to the number of non-vent patients *L_t_*. We model vent-starts 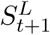 as a random variable that follows binomial distribution with parameters *L_t_* and *p_L_:*

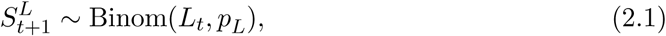

where *p_L_* is a fixed probability that does not change over time. We discuss how to compute probability *p_L_* in Section 2.2.

In addition, on day *t* +1, there will be *A_t_*_+1_ new hospital admissions, and some number of them, 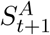, will turn into vent-patients on the same day *t* + 1.

Similarly we assume 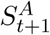 is an independent binomial random variable with parameters *A_t_*_+1_ and *p_A_*:

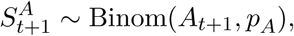

where *p_A_* is a fixed probability that does not change over time. We discuss how to compute parameter *p_A_* in Section 2.3.

Then we estimate the number patients who start the ventilator support on day *t* +1 as

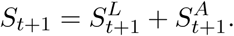

A binomial distribution Binom(*n,p*) can be approximated by Poisson distribution with mean *np* when *n* is large and *np* is moderate. We use this fact to approximate the distribution of the number of vent-start patients as Poisson distribution with mean *p_L_L_t_* + *p_A_A_t_*_+1_:

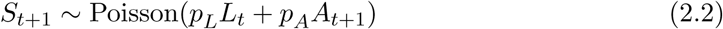

In the following sections we propose a method of estimating parameters *p_L_* and *p_A_*.

### 2.1 Patient types

Each patient is assigned to one of the three patient types: never-vent patient (type 1), vent-cure patient (type 2), and vent-die patient (type 3). The type of a patient is not observable, but does *not* change over time.

We assume that the probability distribution *(p*_1_, *p*_2_, *p*_3_) for a patient to belong to one of these types is known (exogenously, meaning that they do not depend on the congestion levels in hospitals; of course, overly congested hospitals increase death rate.)

We assume that the probability of a type 2 patient becoming a ventilated patient tonight is *q*_2_. Similarly, we use *q*_3_ to denote the probability for a type 3 patient to become a ditto patient tonight. To estimate *q_i_* for *i* = 2, 3, we note that

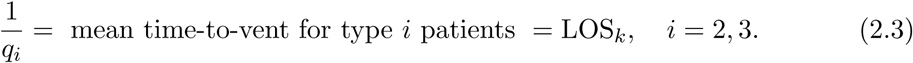

A patient cannot change type; a unknown type will eventually be revealed.

### 2.2 Conversion from *L_t_*

Among *L_t_* patients who are not on vent yet on day *t*, we need to separate them into three types: 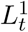, 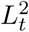, and 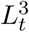.

Motivated by Theorem 1 of [3] in the setting of *M_t_*/*G*/∞ queues, one expects that *L^k^* (*t*) is “close” to a Poisson random variable with mean that is proportional to

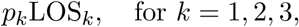

where LOS_1_ is the average length-of-stay for those patients (type 1) who never need a vent, LOS_2_ = 1/*q*_2_ and LOS_3_ = 1/*q*_3_ are average time-to-vent for type 2 and type 3 patients, respectively. Thus, we propose

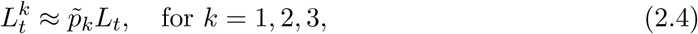

where

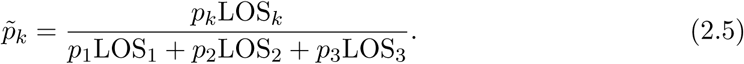

Now we can estimate parameter *p_L_* in (2.2) as

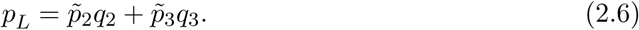

We expect (2.4) can be properly formulated as a functional strong-law-of-large-numbers: for any *t* > 0,

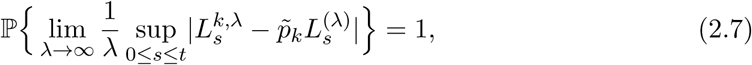

where λ > 0 is a scaling parameter representing the “size” of the hospital. Limits in (2.7) is also known as fluid limits as the “market size” λ goes to infinite. See, for example [4, 5], for a discussion of “large-capacity” scaling. The limit in (2.7) exhibits one form of “state space collapse”, a term coined by [7], meaning that the three-dimensional process “(*L*^1^(*t*), *L*^2^(*t*), *L*^3^(*t*)), *t* ≥ 0} is a deterministic multiple of the one-dimensional process {*L*(*t*), *t* ≥ 0} when the “market size” λ is large.

### 2.3 Conversion from *A_t_*_+1_

We classify the *A_t_*_+1_ admissions on day *t* + 1 by patient type. Thus,

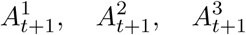

are the number of admitted type 1,2, and 3 patients on day *t* + 1, respectively. Following our assumption,

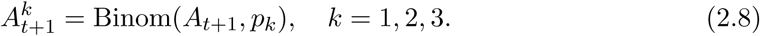

where *p*_1_, *p*_2_, *p*_3_ are given in (1.1). Among 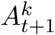 type *k* patients admitted on day *t* + 1,

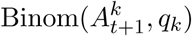

will turn into type *k* vent-patient by the end of day *t* + 1, *k* = 2,3. Thus, among *A_t_*_+1_ admissions on day *t* + 1,

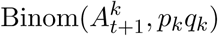

will turn into type *k* vent-patient by the end of day *t* + 1, *k* = 2, 3. Therefore, we can estimate parameter *p_A_* in (2.2) as

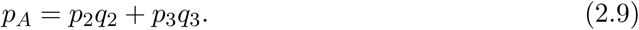

## 3 Ordering and returning ventilators

As before, we assume we are at the end of day *t*. In this section we propose a method that provides the recommended number of ventilators *V_t_*_+1_ to order or return. This tool is intended to ensure that the medical facilities have enough ventilators on day *t* + 1. The tool can also be used to detect surplus of ventilators on day *t* + 1, so that they can be returned to a stockpile or be transported to other locations that require them.

The number of free and ready-to-use ventilators on day *t* + 1 has to to meet the demand of vent-start patients with probability close to 1. According to equation (1.4), number of vent-starts *S_t_*_+1_ follows Poisson distribution with mean 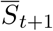. We denote *U_t_*_+1_ the upper confidence bound on the vent-starts on day *t* + 1 as defined in (1.5).

We also recommend to have a *safety-stock* pile with *G* ventilators on the spot that might be used in emergency, unforeseen situations. The size of the safety-stock pile is a hyperparameter and should be determined by a hospital manager who takes into account availability of vent storage facilities, vents delivery speed, etc.

Let *R_t_* be the number of free and ready-to-use ventilators on day *t* at 8pm. Then we recommend to order

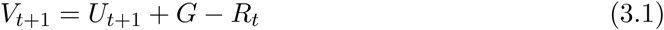

ventilators.

If number *V_t_*_+1_ is negative the hospital can return |*V_t_*_+1_| ventilators back to the federal stockpile.

## 4 Appendix

In this appendix, we test the accuracy of our predictions of vent-starts: mean in (1.3) and upper confidence level *U* in (1.5). We could not find data that included the daily statistics for vent-start patients. In order to test accuracy of vent-starts prediction and efficacy of the proposed ordering policy, we use the simulation model described in Section 4.1. We also test the effectiveness of the policy for ordering and returning ventilators.

### 4.1 Simulation model

To verify the effectiveness of our proposed prediction model for vent-starts in a real hospital setting, we need to know the number of new patients on vents each day. We could not get access to this information even though many cities including New York City (NYC) publicize some related hospitalization data for Covid-19 patients. Therefore, we created a simulation model that would generate the number of daily vent-starts and daily hospital census numbers. Our simulation model uses actual NYC daily admissions and is calibrated so that it matches the NYC daily hospital census. We demonstrate that our predicted vent-starts, using (1.3) and (1.5), matches the vent-starts outputted from the simulation model.

The simulation model takes 3 inputs:

1. Number of hospitalized patients who are not on ventilators on day *t* = 1.
2. Number of hospitalized patients who are on ventilators on day *t* = 1.
3. New admissions per day (series) for each day.

Every new patient is independently sampled as type 1, 2 or 3 with probability *p*_1_, *p*_2_, and *p*_3_ respectively, see (1.1). Then, their patient journey (days in hospital, days-to-vent, and days on a ventilator) is independently sampled from the following distributions depending on their type:

- Geom(LOS_1_), LOS for never-vent patients,
- Geom(LOS_2_) and Geom(LOS_3_) days-to-vent for type 2 and 3 patients,
- 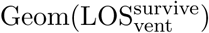 and 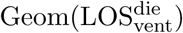 days-on-vent for type 2 and 3 patients,

where LOS_1_, LOS_2_, LOS_3_ are defined in Section 1, 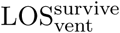 is the average time on ventilator support for type 2 patients before recovery, 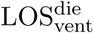 is the average time on ventilator support for type 3 patients before passing away.

We assume that it is impossible to separate type 2 and type 3 patients based on the time from hospitalization to the vent support. Therefore, we assume that ‘time-to-vent’ follows the same distribution for both types of patients:

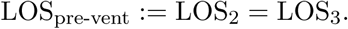

For patients that are already in hospital at the beginning of the simulation, either on ventilators or not, we make the following assumptions. If they are in the hospital but not on ventilators, they are of type *k* with probability 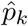, *k* = 1,2,3, and their patient journey is sampled from the same distribution as a new patient, where

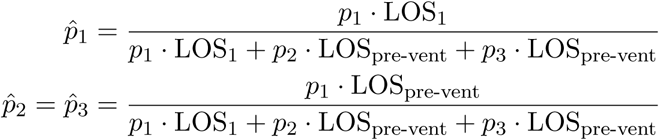

are the same one in (2.5). For patients that are already on ventilators when the simulation begins, we use a formula similar to (2.5) to determine their patient type. Specifically, they are a patient of type *k* with probability 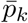, *k* = 2, 3, where

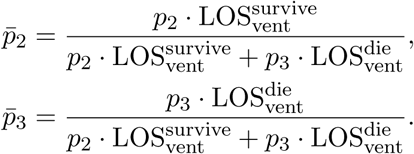

Their length of stay is a geometric random variable with mean 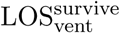 if they are type 2 or 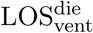 if they are type 3. We also assume they are immediately discharged after their duration on a ventilator ends.

Our simulation model demonstrates high accuracy when predicting the number of patients on ventilators in New York City using the parameters listed in Section 4.2. Figure 1 shows the expected number of patients on ventilators (red dots) and the expected range according to our simulation model (red bars). The range is the maximum and minimum number of ventilators required on a specific day across 100 runs of the simulation. The green triangles represent the real number of patients on ventilators.

**Figure 1:**
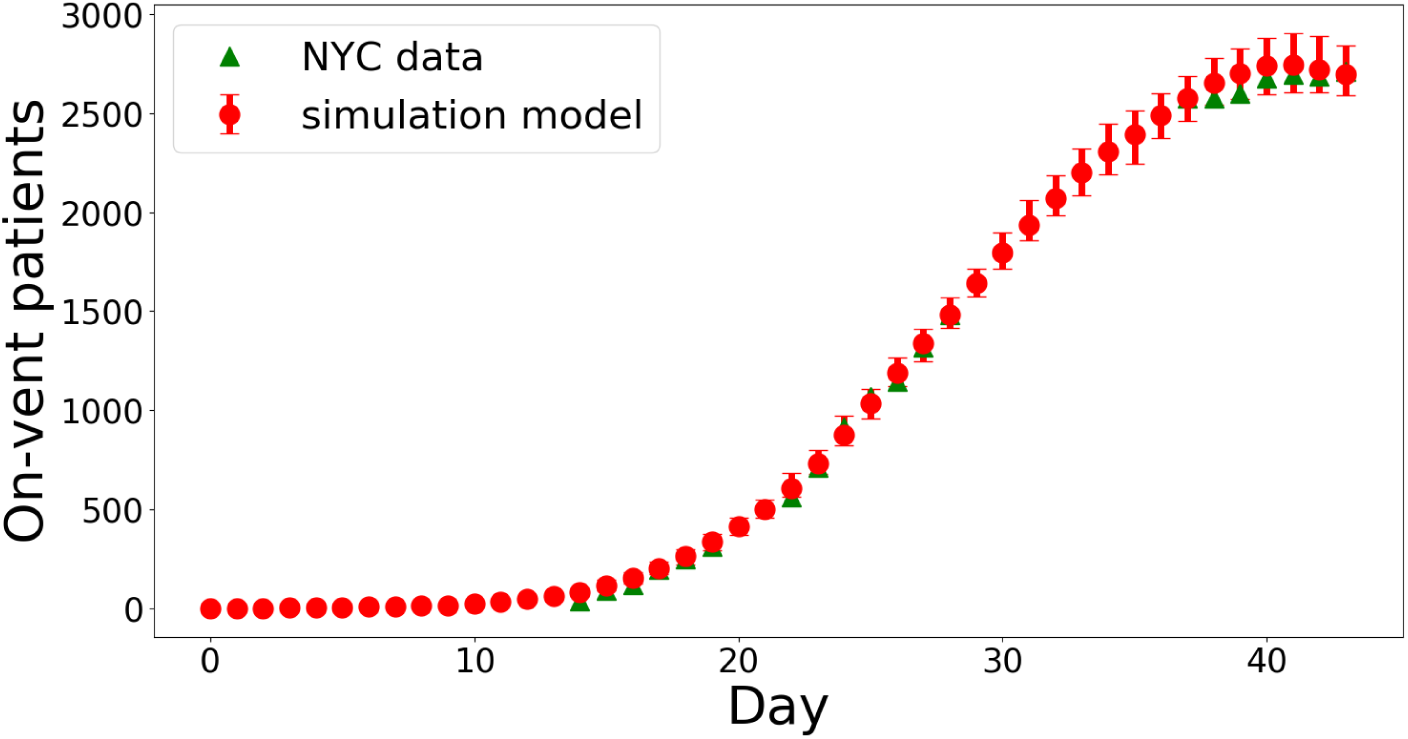
Comparison of the number of patients on ventilators. NYC data is missing on many days.

### 4.2 Simulation Parameters

The parameters for patient type used in the simulation are *p*_1_ = 0.7, *p*_2_ = 0.06, *p*_3_ = 0.24. These numbers are from the CDC website [6] and news reports which said that 80% of all ventilator patients in NYC died [2]. The parameters for length-of-stay used in the simulation are LOS_1_ = 10, LOS_pre-vent_ = 4.8, 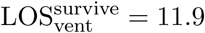, 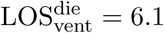. The LOS_1_ parameter is from the CDC website for Covid-19 [6] while the other 3 parameters were tuned using the total number of people on ventilators in NYC between March 16th - 21st and March 24th - 30th. The parameters were tuned using Bayesian optimization and the loss function was mean square error over 50 scenarios.

### 4.3 Numerical experiments

As an input to the simulation model we use real daily admissions of hospitalized patients with COVID-19 at New York City from March 3 to April 14. We assume that there were no hospitalized patients with COVID-19 before March 3 and we set initial number of hospitalized patients to zero for the simulation model run. The model parameters are specified in Section 4.2. In Figure 2 we show the number of on-vent and not-on-vent patients that stay in the hospital according to a simulation run.

**Figure 2:**
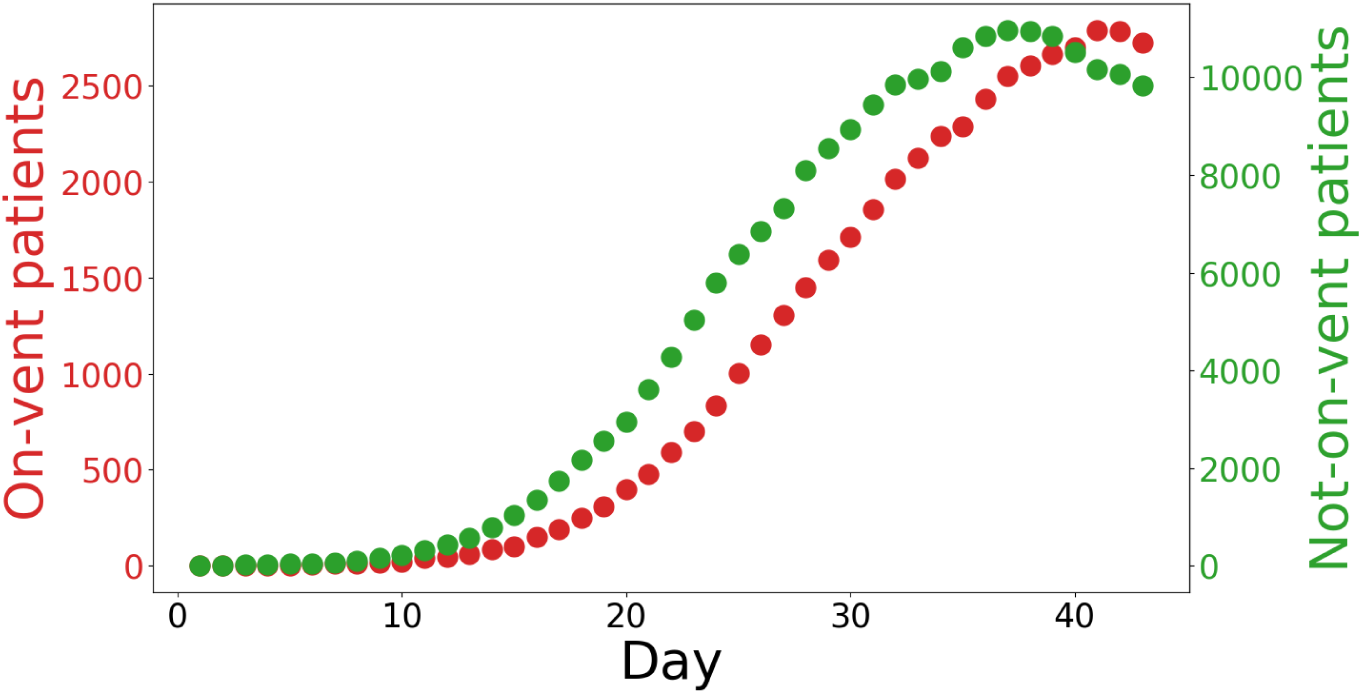
The number of on-vent patients and the number of not-on-vent-patents from the simulation model. The simulation output is from one simulation run.

From the same simulated trajectory we can get daily information about vent-starts. In Figure 3 we plot these vent-starts in green triangles. We use these green triangles as benchmarks to test the accuracy of our prediction formulas (1.3) and (1.5). To recapitulate the prediction procedure described in Section 1, at the end of day *t* we observe the number of hospitalized patients *L_t_* and the historical admissions including *A_t_*, the admission on day *t*. Using the historial admissions, one can predict the number of new admissions *A_t_*_+1_ on day *t* + 1 using a time series model described in Section 4.4. Given *L_t_* and *A_t_*_+1_, one predicts *S_t_*_+1_ the number of vent-start patient for the next day via Poisson model (1.4). In Figure 3 red circles show the expected number of vent-starts, 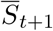, computed by equation (1.3) on each day *t* + 1, *t* = 0,…, 42. The top of the red bars correspond to *U_t_*, the upper confidence bound on the vent-starts computed from (1.5).

**Figure 3:**
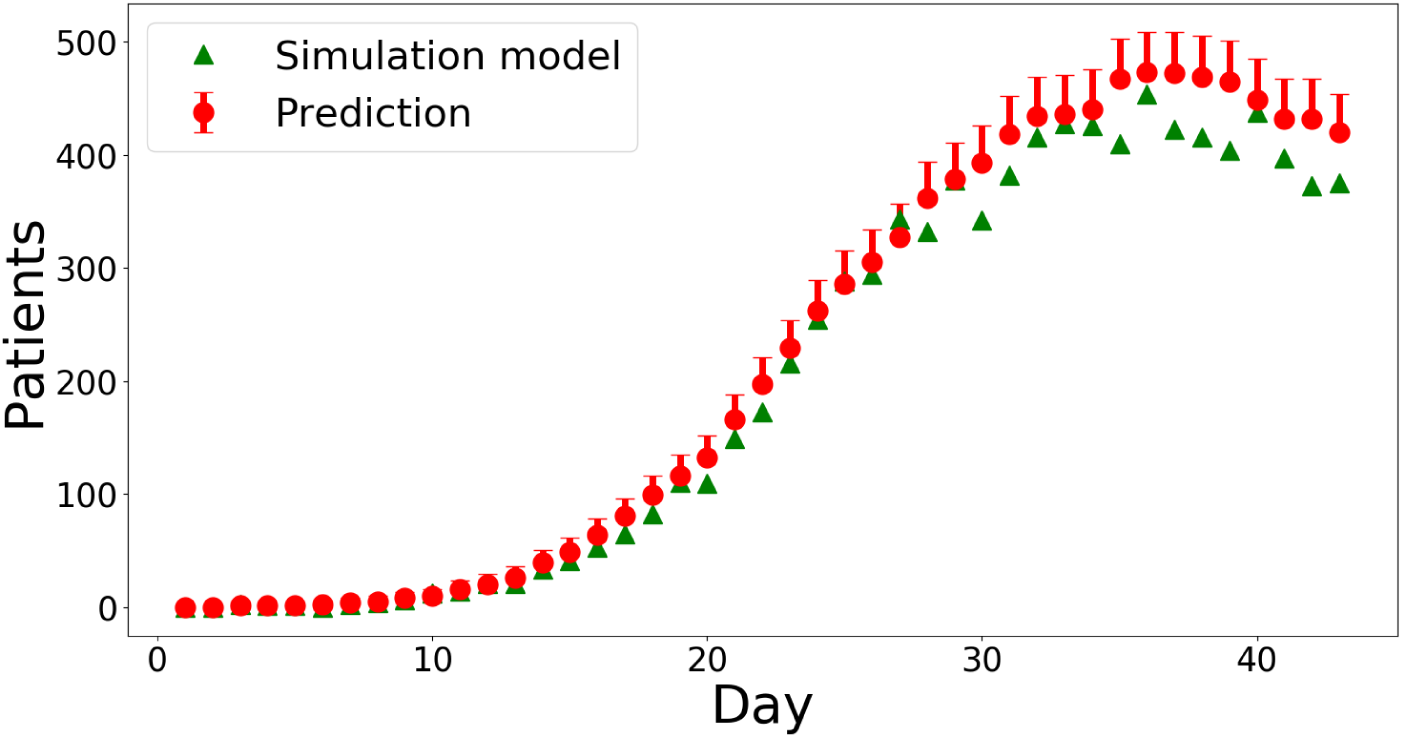
Prediction accuracy of the number of vent-start patients. *The green triangles* shows the number of vent-start patients according to the simulated trajectory, *the red circles* – expected number of the vent-starts according to (1.3), *the top of the red error bar* – upper confidence bound on the vent-starts according to (1.5).

Next we test the ordering policy proposed in Section 3. Using the prediction of the vent-starts *U_t_*_+1_ on day *t* + 1, we either order *V_t_*_+1_ ventilators from a central stockpile that need to be delivered by day *t* + 1 when *V_t_*_+1_ > 0 or return | *V_t_*_+1_ | ventilators to the central stockpile when *V_t_*_+1_ < 0, where *V_t_*_+1_ is computed according to formula (3.1). We assume that if *V_t_*_+1_ < 0, the hospital sends back | *V_t_*_+1_ | ventilators in the morning of day *t* + 1. In Figure 4a we provide the number of ordered/returned ventilators on each day. We set the safety stock level to be equal to *G* = 10.

**Figure 4:**
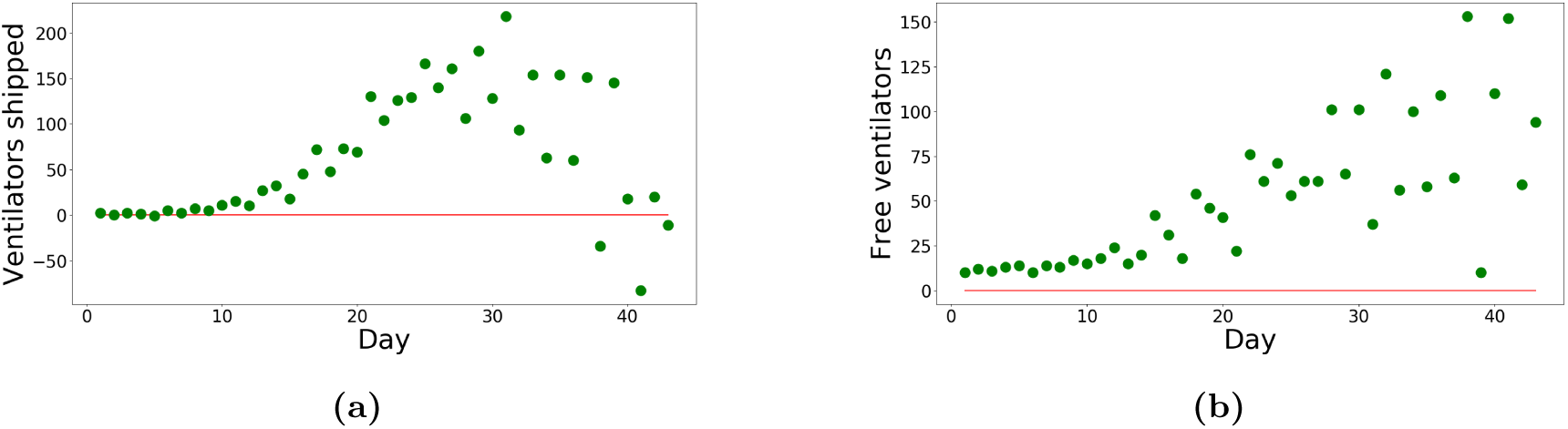
Efficiency of the recommend policy. (a) The daily number of ventilators ordered/returned according to the policy, (b) surplus of ventilators at the end of each day.

In Figure 4b we show the number of unused ventilators at the end of each day that are possessed by the hospital. We note that the plot demonstrates that the hospital always has enough ventilators to satisfy the demand from patients who need ventilator support. On the other hand, the hospital is not oversupplied from the central stockpile.

We run 100 independent simulations runs to test the robustness of the proposed policy. In Figure 5 we show minimum, average and maximum number of free (ready-to-use) ventilators observed during each independent simulation run. We observe that the hospital could satisfy the demand of ventilators from vent-start patients and, at the same time, did not accumulate too many free ventilators by the end of each day.

**Figure 5:**
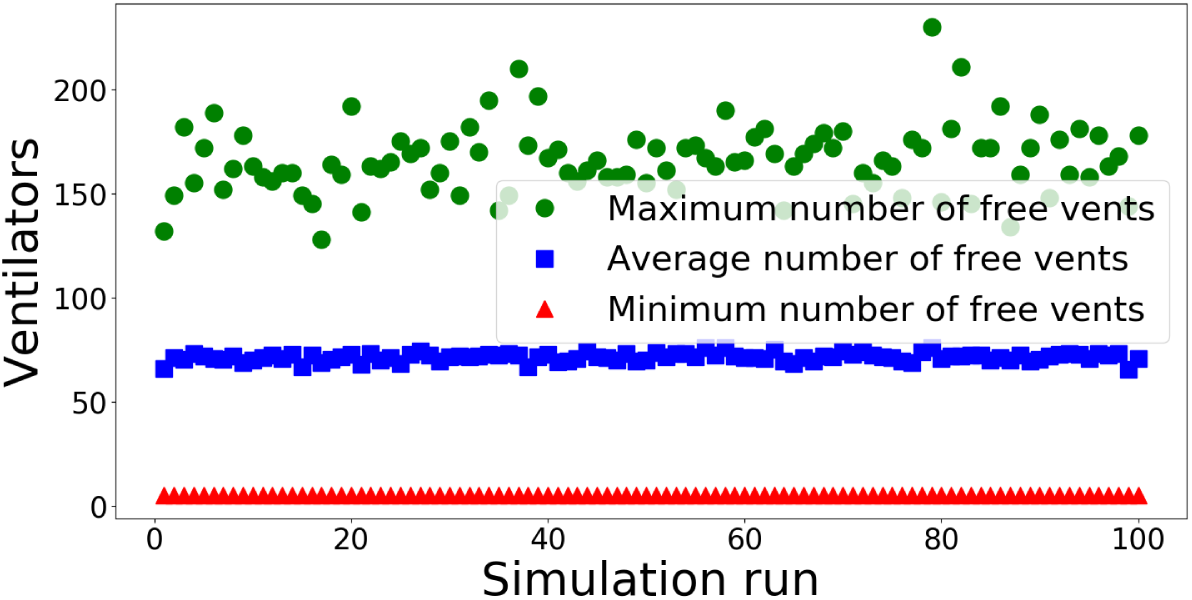
Robustness of the policy for ordering and returning ventilators. No patients were denied of ventilation, no excessive inventory of ventilators kept at the hospital.

### 4.4 Predicting the number of admissions next day

Since we make a short term prediction (tomorrow) of the number of hospital admissions, we use a standard time series algorithm to make this prediction, assuming historical daily admission numbers are available. We adopt the algorithm ARIMA(*p*, *d, q*) in [1], where parameters *p, d*, *q* are tuned based on the input of historical data. It has been well developed in many library packages. For example, `Python` has a library function that implements this algorithm with automatic tuning. We set the algorithm to be adaptive, meaning that every day, as a new data point becomes available, the parameters *p, d, q* are re-tuned and the algorithm gives a new prediction for the next day. After fitting the ARIMA model on the historical admission data, the prediction for the next day’s hospital admissions closely matches the observed value. This suggests that prediction of the ARIMA model could be a reliable input for anticipated hospital daily admission next day. Figure 6 is an example of a 3-day prediction of NYC hospital daily admissions, with 95% confidence interval.

**Figure 6:**
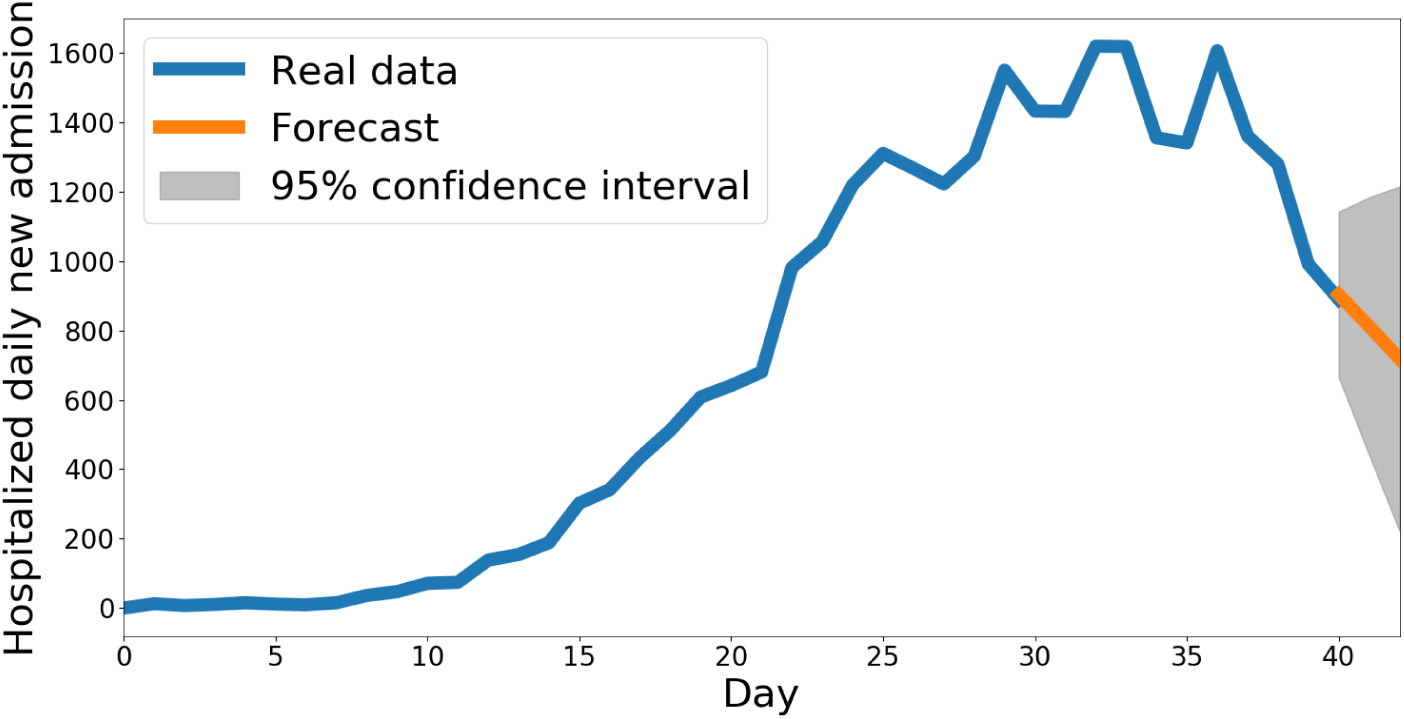
New York City Hospitalized Daily New Admission

## Data Availability

The data is available through a web interface at covidvent.github.io

http://covidvent.github.io

